# Design considerations for incorporating serological monitoring into trachoma prevalence surveys

**DOI:** 10.1101/2025.09.05.25334960

**Authors:** Everlyn Kamau, Katherine Gass, Emma M. Harding-Esch, Martha Idalí Saboyá Díaz, Pearl Anne Ante-Testard, Amir B. Kello, Robin L. Bailey, Kimberly Fornace, Anthony W. Solomon, Scott D. Nash, Benjamin F. Arnold, Global Trachoma Serosurveillance Study Group

## Abstract

Serology is increasingly used to monitor infection transmission and disease elimination. Embedding dried blood spot collection in trachoma surveys allows transmission intensity inference through population-level seroconversion rate (SCR) estimation, but no formal assessment of required sample size has previously been published. Using data from 40 prevalence surveys, we estimated intra-cluster correlation coefficient, a key design parameter, and assessed survey design considerations for incorporating serological monitoring. Design scenarios focused on proposed operational SCR thresholds (2.2 [no action needed] and 4.5 [action needed] per 100 child years) for interpretation of serological data in low-transmission and post-elimination settings. We evaluated SCR estimation in 42 two-stage designs by calculating precision (confidence interval width around an SCR value) and power (the measure of deviation of suggested thresholds to the SCR value). When the SCR is ≤1.5 and >5.7 per 100 person-years, sample sizes between 300-2000 allowed good precision of SCR estimation. We anticipate a standard trachoma prevalence survey with 30 clusters and 30 households per cluster should provide ≥80% power to determine that an SCR ≤1.2 falls below a 2.2 threshold and SCR ≥5.7 falls above a 4.5 threshold. Our results support estimation of serological data via the recommended population-based survey design for trachoma surveillance.

## Introduction

Trachoma is a blinding disease caused by repeated conjunctival infection with *Chlamydia trachomatis* (*Ct*). Prevalence estimates of the ocular sign “trachomatous inflammation—follicular” (TF) among children aged 1–9 years are used to guide intervention planning including implementation of antibiotic mass drug administration (MDA) ^1^. The World Health Assembly has targeted global elimination of trachoma as a public health problem by 2030, with elimination defined at the level of the evaluation unit (EU; please see below) as: a prevalence of TF <5% in 1–9-year-olds in each formerly-endemic EU, a prevalence of trachomatous trichiasis (TT) unknown to the health system of <0.2% in ≥15-year-olds in each formerly-endemic EU, and a system in place to identify and manage incident TT ^2^.

The WHO-recommended population-based trachoma prevalence survey design estimates TF prevalence within the usual administrative units for healthcare management, generally with populations of 100000–250000 people; otherwise referred to as EUs. Prevalence estimates are EU-specific, and EUs are the level of programmatic decision-making. WHO’s recommended sampling approach and field methodology incorporates two-stage cluster sampling within EUs, which involves (i) selection of villages, census enumeration areas or the local equivalent as primary sampling units (“clusters”); and (ii) selection of households within a cluster as the secondary sampling units ^3^. First- and second-stage sampling use methods that equalize, as far as practicable, the probability that any given individual resident aged ≥ 1 year in the EU will be invited to participate. A minimum of 20 clusters, and a maximum of 30, is recommended per EU ^3^. To maximize efficiencies, it is recommended that the number of households per cluster is set as the number that a team (one grader plus one recorder) can comfortably see in a single day, which will vary based on distances between households and the geography of the intervening terrain ^3^. In each selected household, a certified grader examines each consenting and assenting resident aged ≥ 1 year for the clinical signs of TT, TF and trachomatous inflammation—intense (TI) ^3^. TI is assessed so that graders do not flag severe conjunctival inflammation as TF if the definition of TF is not also met ^4^. Tropical Data supports health ministries to conduct trachoma prevalence surveys in a standardized way, adhering to WHO recommendations, and with quality assurance and control at every stage of the survey process. Further details on cluster and household selection procedures and survey implementation can be found in WHO and Tropical Data publications ^3,5^.

Presence of detectable *Ct* DNA at ocular sites (often referred to as *Ct* infection) and serological markers are two common biological measures of trachoma transmission intensity. In EUs that implement MDA for trachoma, the prevalence of TF may remain elevated even after MDA has successfully cleared current conjunctival infection episodes ^1^. The relationship between TF and the biological measures of infection thus weakens in post-MDA settings ^6^, and so DNA detection and/or serology may complement TF to guide programmatic decision-making post-MDA. Measurement of anti-Pgp3 IgG antibodies (serology) has become valuable for quantifying *Ct* transmission intensity in routine trachoma population-based surveys ^7^. The sensitivity of IgG measurements may help detect *Ct* recrudescence as countries approach elimination and after validation of elimination as a public health problem ^6,8^, but this way of assessing populations has not yet been formally recommended by the World Health Organization (WHO).

Here, we assessed two-stage sampling designs by evaluating statistical uncertainty around seroconversion rate (SCR) estimates among 1–5-year-old children. Limiting the age range to pre-school age children ensures that serology reflects recent infections and avoids the potential contribution of sexually transmitted *Ct* ^8^. Unlike seroprevalence, the SCR implicitly adjusts for age and, by quantifying incident seroconversions from ages one to five years, captures postnatal (rather than congenital) *Ct* exposure ^7^. We evaluated sampling designs for estimating SCR among 1–5-year-olds within the standard EU-level framework for TF surveillance and used simulations to assess designs that would optimize precision and power around target SCR thresholds.

## Methods

### Serological surveys

We gathered serology data on IgG antibody responses to the *Ct* antigen Pgp3 obtained from 40 surveys in the Global Trachoma Serology Data Repository ^9^ (Supplementary Materials). The surveys spanning from elimination to hyperendemic settings, ranged in Pgp3 seroprevalence from 0.5% (95% confidence interval [CI], 0%-1.1%) to 50.7% (44.5%-56.2%), and in SCR from 0.2 (0-0.7) to 14.3 (10.6-19.2) per 100 person-years (**Tables S1 and S2**). The median (range) number of clusters among the 40 surveys was 24 (12–50), while the median sample size of 1–5-year-olds was 623 (166–1301). We only included surveys conducted using WHO-recommended methodologies ^3^, with the addition of dried blood spot collection. We categorized surveys into three groups based on proposed programmatic actions: (i) ‘*action needed*’ - those believed to be likely to experience development of disease sequelae and blindness from trachoma in the absence of effective interventions, (ii) ‘*action not needed*’ - those believed to be unlikely to have sufficiently intense and sustained conjunctival *Ct* transmission to lead to blindness, and thus no contemporary justification for population-level interventions such as MDA, and (iii) ‘*unclassified*’ – those believed to fall between ‘action needed’ and ‘action not needed’; these EUs might be in transition or have unusual epidemiology (for example, high TF prevalence but low or absent conjunctival *Ct* transmission intensity) ^8^. The reference “the development of disease sequelae and blindness” relates to any sequela or blindness and is not in relation to a specific threshold or incidence rate of disease or blindness.

### Estimation of Intracluster Correlation Coefficient

We estimated the intracluster correlation coefficient (ICC) (**Supplementary Material**) from serological surveys and used this parameter as a surrogate for spatial heterogeneity of *Ct* transmission. We used a similar estimation approach as a previous study that reported the ICC for TF from 261 trachoma surveys in Ethiopia, Mozambique, and Nigeria ^10^.

### Survey design and sample size calculations

We simulated two-stage survey designs of varying sample size, *n* = *k* (number of clusters) × *m* (number of households per cluster), where *k* = {20,25,30,35,40,45,50} and *m* = {15,20,25,30,35,40}. The current recommended designs for estimating TF prevalence, which sample approximately 20–30 clusters per EU and 20–35 households per cluster ^13,14^, are relatively similar to our simulations. While 30 households per cluster is a reasonable maximum for a survey team to complete in a day, we extended the simulations to *k* to 50 and *m* to 40 to see if the tradeoffs in improved precision would merit expanding the size of survey teams or their time in the field. The scenarios here were chosen to represent a reasonable and attainable range of households in various settings, assuming that (i) a single household would have at least one child aged 1-5 years and, (ii) to ensure repeatability and simplify logistics, a survey team can consistently visit at least 30 households per day.

For each design, we calculated the effective sample size, *n_eff_* = *n* / design effect (**Supplementary Materials**). For ease of interpretation in terms of SCR per 100 person-years, which may be used to guide trachoma programmatic decision-making, we used a linear relationship to convert seroprevalence (π) to the SCR: *scr* = (π * 100)0.36 (more details in ^8^). We set π between 0.5% and 30%, which represents the breadth of possible seroprevalence estimates in the majority of the 40 surveys (39/40, **Table S1**). Additionally, for each design of size *n_eff_* and π, we evaluated power to detect a difference from two proposed SCR thresholds, 2.2 and 4.5, which correspond to 90% probability of *no* action needed (<2.2) and action needed (≥4.5) after halting MDA or during surveillance after WHO validation of elimination ^8^. We mostly focus on the 2.2 threshold with the idea that decision-making in trachoma programs during the surveillance phase would be more geared towards confidence that *no* further action or intervention is needed.

We used the width of the 95% CI around a given SCR to compare designs by evaluating precision of SCR estimates relative to the 2.2 per 100 person-years threshold. We considered power as the probability of correctly rejecting the null hypothesis that a sample seroprevalence estimate does not statistically differ from a predefined value; more specifically, the probability of detecting a statistically significant difference in SCR if a true difference exists. The power analyses were done using the *propTestPower* function in the package EnvStats ^15^ based on two-tailed, one-sample hypothesis testing, *H*_0_: *π* = *π*_0_ versus *π* ≠ *π*_0_, where *π* denotes seroprevalence, and *π*_0_ is a fixed target value representing the SCR threshold.

For all simulations, we used two ICC values, *ρ*= {0.002, 0.01} to represent low (*ρ*=0.002) and high (*ρ*=0.01) spatial heterogeneity of anti-Pgp3 IgG seroprevalence in the context of near elimination thresholds. These two values represent the mean and upper 95% CI of the pooled ICC value for EUs in the *action not needed* category (see below). Conceptually, in the context of the *action not needed* category, these two ICC values could align with locations where interventions had been successful in curbing transmission (low, *ρ*=0.002) versus those with strong candidacy for successfully curbing transmission with concerted efforts (high, *ρ*=0.01). In the latter case (*ρ*=0.01), this could be a district where one village / cluster has high transmission but EU-level seroprevalence remains below the threshold to warrant EU-level treatment. So, the decision might involve efforts to curb infections in that one village if there is concern this could drive transmission further.

## Results

In the *action not needed* category, ICC estimates were homogenous and below 0.1 in all but two surveys, and always in the range of 0–0.16 (**Figure 1**, **Table S2**). ICC estimates were higher and more varied among EUs categorized as *unclassified* (range, 0-0.31) and *action needed* (range, 0.03–0.42), consistent with the observed cluster-level heterogeneity in seroprevalence (**Figure 1**). Overall, there was an overlap in ICC distributions by endemicity categories and ICC was not discernably linearly correlated with seroprevalence. The pooled ICC estimate for Pgp3 seroprevalence across all 40 surveys was 0.007 (95% CI, 0.002–0.02), while in the *action not needed* category it was 0.002 (0.0003–0.01). In the *action needed* category, the pooled ICC estimate was 0.13 (0.05–0.32).

**Figure 1.**
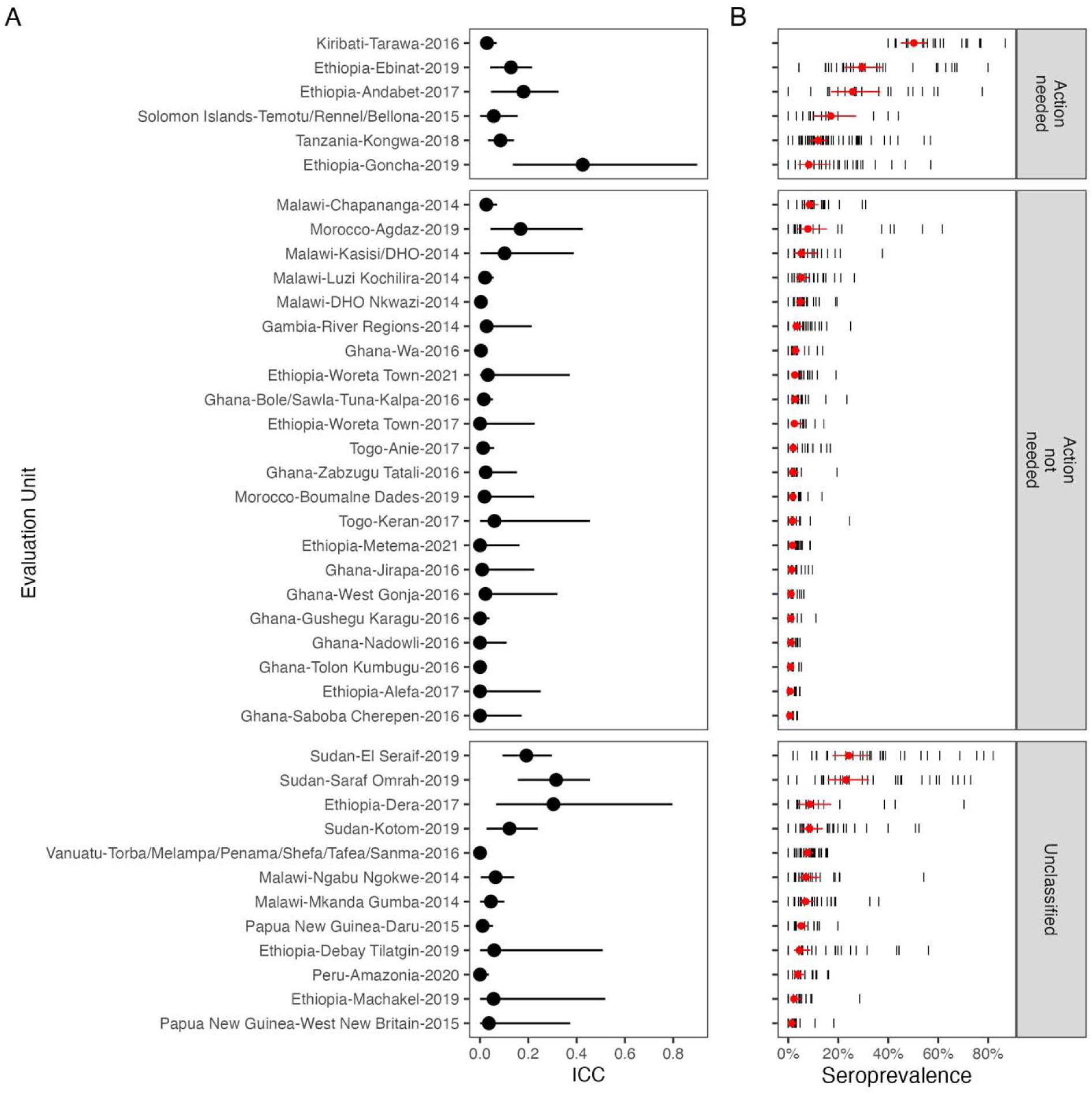
Intracluster correlation coefficient (ICC) estimates for 40 evaluation units (EU) (A) and their corresponding Pgp3 seroprevalence estimates (B). The individual black vertical lines in Panel B show cluster-level seroprevalence while the red circles and horizontal lines indicate the EU-level seroprevalence with 95% confidence interval (CI). The pooled ICC estimate across all surveys was 0.007 (95% CI, 0.002–0.02). The pooled ICC in the *Action not needed* category was 0.002 (0.0003–0.01) and in the *Action needed* category was 0.13 (0.05–0.32). The data (mean and 95% CI) are sorted by seroprevalence and grouped by category of programmatic response (see Methods for definitions of the categories). Alt text: Intracluster correlation coefficient and seroprevalence estimates for 40 trachoma prevalence surveys.

### Precision and power of two-stage survey designs

Our results suggest that for EU-level SCR values ≤1.5 and ≥3, all but the smallest surveys will have 95% CI that exclude the 2.2 threshold (**Figure 2**). If the SCR is 1 per 100 person-years, all 42 design scenarios would yield upper bounds of the 95% CI that are below the 2.2 per 100 person-years SCR threshold (**Figure 2**). For an SCR of 1.5 per 100 person-years, smaller sample sizes (*k* ≤ 25 and *m* ≤ 20) would yield an SCR with an upper bound of 95% CI above the 2.2 threshold, and this is more evident when assuming higher heterogeneity of seroprevalence between clusters (ICC = 0.01). For an SCR of 2 per 100 person-years, the upper bounds of the 95% CI are above the 2.2 threshold in all design scenarios, and when the SCR is 2.5 per 100 person-years, then all the lower bounds fall below the 2.2 threshold (which generally applies to SCR <2.5) (**Figure 2**). Smaller and medium samples (*k* ≤ 40 and *m* ≤ 20) in an EU with an SCR of 3 per 100 person-years would result in the lower bound of the 95% CI being below the 2.2 threshold. **Appendix S1** includes detailed estimates of precision (95% CI width) across a range of study designs and parameters.

**Figure 2.**
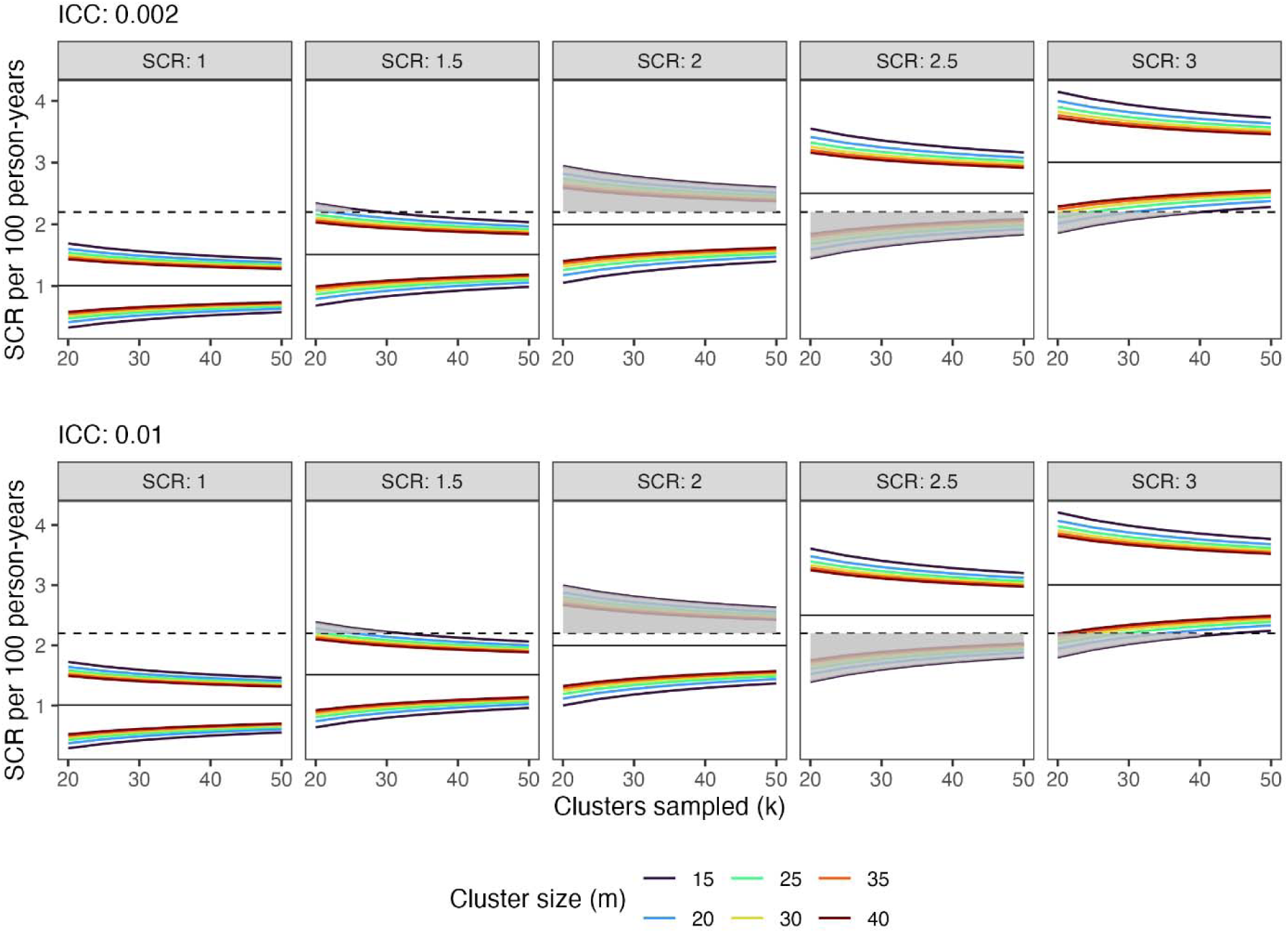
Precision of Pgp3 seroconversion rate (SCR) estimation across various designs and example population SCRs relative to a threshold. Line colors depict cluster sizes (or number of children per cluster) and shaded regions depict the region of SCR above or below an SCR of 2.2 per 100 person-years. An SCR of 2.2 corresponds to 90% probability of *no* further need for trachoma-related action based on a previous analysis. The two intracluster correlation coefficient (ICC) values were adopted from estimates in the populations in Figure 1, and SCR values ranging from 1 to 3 represent the SCR for populations near or post elimination. **Appendix S1** includes detailed estimates presented in this figure across the full range of design scenarios. Alt text: Precision of SCR estimation relative to a threshold of SCR 2.2 per 100 person-years

To complement the precision analysis, we evaluated designs that would have ≥80% power at 95% significance level relative to the 2.2 per 100 person-years SCR threshold. We observed, as expected, that a design’s power to determine if SCR falls above or below a threshold declines as the SCR approaches the threshold. This challenge is inherent to any decision-making rule with respect to a cutoff. At ≥80% power and relative to the SCR = 2.2, and at ICC of 0.002, the largest survey (*k*=50 × *m*=40) would be powered to detect a true difference when the EU-level SCR is <1.6 and >2.7, however, such a large survey would be expensive and might be infeasible practically. The smallest scheme (*k*=20 × *m*=15) would be powered to detect a true difference when the EU-level SCR is <0.9 and >3.7 (**Figure 3**). Across the 42 design scenarios, the probability of detecting a true difference from the threshold SCR of 2.2 is 80% when the EU-level SCR falls in the range of 0.9–1.6 and 2.7–3.7 per 100 person-years. Smaller sample sizes (range, 300–1200) would be restrictive such that power falls below 80% for a wider region around the 2.2 threshold (0.9–2.9) compared to larger samples (1225–2000) where power falls below 80% in a narrower region (1.6–2.7) (**Figure 3**).

**Figure 3.**
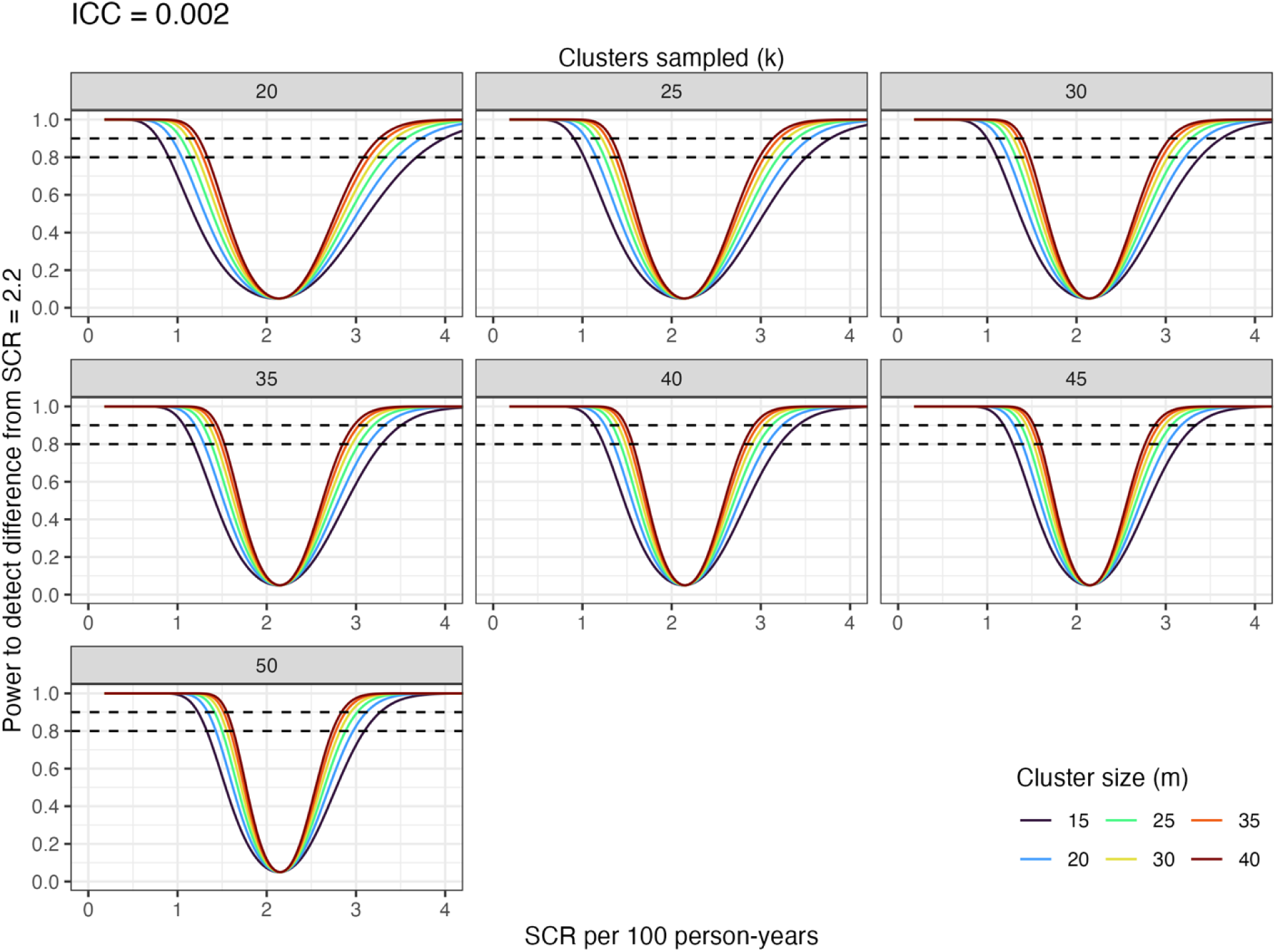
Estimated power of various designs to distinguish Pgp3 seroconversion rates (SCR) relative to an SCR threshold of 2.2 per 100 person-years. An SCR of 2.2 corresponds to 90% probability of *no* further need for trachoma-related action based on a previous analysis. The different colored lines depict cluster sizes (or number of households per cluster) and the dashed lines indicate 80% and 90% power levels. The analytical estimates here assume a low intracluster heterogeneity (intracluster correlation coefficient, ICC = 0.002) estimated in settings with no action needed and SCR values represent the breadth of possible underlying SCR as estimated in different populations in the 40 surveys analyzed here. Alt text: Power of SCR estimation relative to a threshold of SCR 2.2 per 100 person-years

When assuming higher between-cluster heterogeneity (ICC = 0.01), the largest survey (*k*=50 × *m*=40) would be powered to detect a true difference when the EU-level SCR is <1.5 and >2.8; while the smallest scheme (*k*=20 × *m*=15) would be powered to detect a true difference when the EU-level SCR is <0.8 and >3.8 (**Figure S1**). Similarly, this means that across the 42 design scenarios, the probability of detecting a true difference from the threshold SCR of 2.2 is 80% when the EU-level SCR falls in the range of 0.8–1.5 and 2.8–3.8 per 100 person-years. For survey designs relative to the SCR threshold of 4.5 and assuming ICC = 0.002, the power level drops below 80% when the EU-level SCR falls in the range of 3.5–5.1 per 100 person-years in the largest survey (*k*=50 × *m*=40) versus 2.5–6.3 in the smallest survey (*k*=20 × *m*=15) (**Figures S2**). With an assumed ICC of 0.01, the two designs (*k*=50 × *m*=40 and *k*=20 × *m*=15) are powered at 80% in the SCR range of 3.5–5.2 and 2.4–6.4, respectively (**Figure S3**).

The *k*=30 by *m*=30 design was previously developed to provide sufficient precision around the TF and TT prevalence estimate and is based on the number of households a survey team can comfortably survey within a single day ^3,5,16^. If each household per cluster has at least one eligible child participant, the number of households per cluster ≍ number of children per cluster. Therefore, *k* clusters × *m* households / cluster ≍ *k* clusters × *m* children / cluster ≈ number of children providing a serum sample. We anticipated that the *k*=30 by *m*=30 design for TF and TT prevalence estimation would yield an equivalent sample of 600 households (≈ 600 children of age 1-5 years), which was close to the median number of 1-5-year-olds in our reference dataset (**Table S1**). That is, if field teams are going out with the recommended design of 30×30 in mind, under ideal circumstances, at least 20 of 30 households per cluster would each have a 1-5-year-old, therefore an equivalent sample of 600 households (≈ 600 1-5-year-olds). So, we focused on detailed estimates for the region of SCR attainable with confidence at 80% to 90% power levels under the *k*=30 by *m*=20 design.

When the spatial heterogeneity is assumed to be low (ρ = 0.002), a survey with the 30×20 scheme has 80% to 90% power to reliably characterize such EUs as having SCR <2.2 per 100 person-years when the SCR is between 1.1–1.2 (light shaded regions, **Figure 4A**). This is very close to the scenario with a higher spatial heterogeneity assumption (ρ = 0.01) (light shaded regions, **Figure 4B**). The assumed level of cluster heterogeneity in SCR (ρ = 0.002 versus 0.01) had minimal impact on precision and power inferences, suggesting that accurate SCR estimates could be made with smaller effective sample sizes, *n_eff_* = *n*/(1 + (*m* - 1) * ρ). So, for the *k*=30 × *m*=20 scheme, an *n_eff_* of ∼504 (= 600 / (1 + (20 - 1) × 0.01)) would have ≥80% power for targeting SCR <1.2 per 100 person-years. This is not substantially different from a larger *n_eff_* of ∼578 (= 600 / (1 + (20 - 1) × 0.002)), which would have ≥80% power for targeting the same level of SCR.

**Figure 4.**
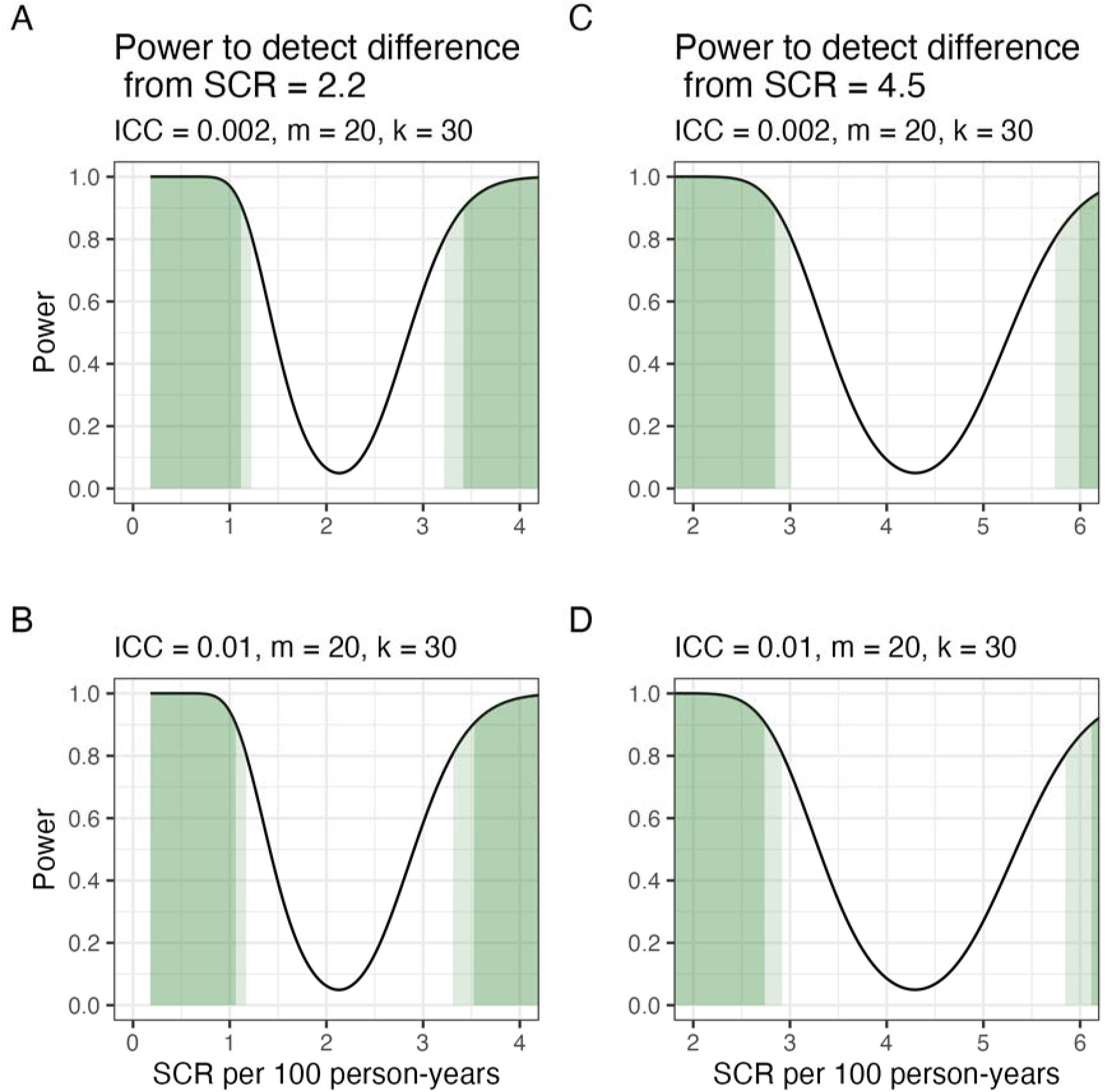
Estimated power of a design with k=30 clusters and m=20 households per cluster to distinguish Pgp3 seroconversion rate (SCR) relative to SCR thresholds of 2.2 and 4.5 per 100 person-years. The 2.2 threshold corresponds to 90% probability of *no* further need for trachoma-related action, while the 4.5 corresponds to 90% probability that further trachoma-related action is needed based on a previous analysis. The lighter green shading depicts the region of SCR detectable at ≥80% power while the darker green depicts ≥90% power. Estimates assume a low (intracluster correlation coefficient, ICC = 0.002) and high (ICC = 0.01) intracluster heterogeneity and the SCR values represent the breadth of possible underlying SCR as estimated in different populations in the reference dataset of 40 surveys analyzed here. Alt text: Power of SCR estimation relative to a threshold of SCR 2.2 per 100 person-years for a sample of 600

The 30×20 scheme would have ≥80% power to detect a true difference from the 4.5 SCR threshold when the EU-level SCR is above 5.7 per 100 person-years. In detail, relative to the 4.5 threshold, a 30×20 survey has 80% to 90% power to confidently detect SCR between 2.8–3.0 and 5.7–6.0 when the spatial heterogeneity is assumed to be low (ρ = 0.002) (light shaded regions, **Figure 4C**). The same design assuming higher spatial heterogeneity (ρ = 0.01) has 80% to 90% power to detect SCR between 2.7–2.9 and 5.8–6.1 per 100 person-years (light shaded regions, **Figure 4D**).

Under an assumed ICC of 0.002, representative of low spatial heterogeneity in populations where no further action is needed, the total sample size required is small and reasonably constant for a fixed SCR (**Table 1**). In the *Action needed* scenarios with an ICC of 0.13, representing the more spatial heterogeneity of anti-Pgp3 IgG seroprevalence, sample size requirements were generally larger when the number of clusters was fixed at k = 20, reflecting the larger cluster sizes and higher design effects under these assumptions. By contrast, when k = 25 or k = 30, the required total sample sizes were far lower than k = 20, consistent with more efficient designs including a larger number of small-sized clusters given the higher ICC (**Table 1**). Survey designs for EUs with SCR between 1.5 and the 2.2 threshold, and between the 4.5 threshold and 7.0 would require ≥30 clusters and >900 households to achieve 80% power (**Table 1**).

**Table 1.** Sample size (product of No. clusters and Cluster size) needed to estimate varying levels of seroconversion rate (SCR) with 80% or more power. The sample sizes would be powered for correct classification into the categories representative of programmatic action. Here, we limit the number of clusters to less than 30 and assume low and high intracluster heterogeneity in seroprevalence (intracluster correlation coefficient, ICC). ICC values of 0.002 and 0.13 were the pooled mean estimates for the two categories.

| Scenario | ICC | SCR per 100 person-years | No. clusters | Cluster size | Design effect | Total sample size |
| --- | --- | --- | --- | --- | --- | --- |
| Action not needed<sup>a</sup> | 0.002 | 1.0 | 20 | 18 | 1.03 | 360 |
|  |  | 1.0 | 25 | 14 | 1.03 | 350 |
|  |  | 1.0 | 30 | 12 | 1.02 | 360 |
|  |  | 1.3 | 20 | 36 | 1.07 | 720 |
|  |  | 1.3 | 25 | 29 | 1.05 | 725 |
|  |  | 1.3 | 30 | 24 | 1.04 | 720 |
|  |  | 1.4 * | 30 | 31 | 1.06 | 930 |
| Action needed<sup>b</sup> | 0.13 | 6.8 * | 30 | 32 | 5.0 | 960 |
|  |  | 7.4 | 20 | 38 | 5.81 | 760 |
|  |  | 7.4 | 25 | 15 | 2.82 | 375 |
|  |  | 7.4 | 30 | 10 | 2.17 | 300 |
|  |  | 7.5 | 20 | 27 | 4.38 | 540 |
|  |  | 7.5 | 25 | 12 | 2.43 | 300 |
|  |  | 7.5 | 30 | 10 | 2.17 | 300 |

<sup>a</sup> Areas believed to be likely to experience development of disease sequelae and blindness from trachoma in the absence of effective interventions. SCR threshold for this category = 2.2 per 100 person-years.
<sup>b</sup> Areas believed to be unlikely to have sufficiently intense and sustained conjunctival *Chlamydia trachomatis* transmission to lead to blindness, and thus no contemporary justification for population-level interventions such as mass drug administration. SCR threshold for this category = 4.5 per 100 person-years.
\* These SCR scenarios would require more than 900 households to achieve ≥80% power to detect true differences from the 2.2 and 4.5 thresholds.

## Discussion

We found that multi-stage designs similar to those currently used to estimate prevalence of clinical signs of trachoma would provide adequate precision for estimating SCR to inform programmatic action. Through two complementary lenses of precision and power informed by empirical ICC estimates for serology, we found that a range of feasible designs will provide clear results for SCR estimates ≤1.5 and ≥ 3 near a stopping threshold of 2.2 per 100 person-years, but if the SCR falls within that range, a survey will provide equivocal results. A design with 30 clusters and 30 households per cluster is a current standard survey recommended to provide sufficient precision for TF and TT prevalence estimates and is designed so that a survey team with a single grader can fully complete one cluster per day ^3,5,16^. This 30×30 design targets 1-9-year-olds, so again assuming that each household has at least one child in that target age group (1-9 years), then it’s likely that 20/30 households per cluster will have at least one child in the age group 1-5 years old. Our analysis suggests that such a design with 30 clusters × 20 households would have favorable power and precision for serology: ≥80% power to determine that an SCR ≤1.2 falls below a 2.2 threshold and SCR ≥5.7 falls above a 4.5 threshold (**Figure 4**).

In surveys designed to assess trachoma elimination through serology that are not necessarily nested within TF and TT prevalence surveys, we show that including up to 30 or even 40 households per cluster would lead to improved precision and power for the SCR estimate, assuming ICCs identified in populations with no further action needed (0.002 to 0.01) and 30 clusters per EU (**Figure 2**, **Figure 3**). Trachoma programs may consider over-sampling children aged 1–5 years within the TF and TT prevalence surveys to improve SCR estimation; however, potential operational implications and risks to the reliability and comparability of TF estimates should be carefully assessed. Any such approach should remain aligned with WHO guidance and standardized protocols.

Estimates from this study pertain to surveys that measure children 1-5-years-old and estimate the SCR using a catalytic model assuming a constant rate over the age range. While this assumption is most suitable in the context of stable endemicity, in post-MDA settings where there has been declining incidence and prevalence a survey of 1-5-year-olds estimates the average recent force infection experienced by children born in the past five years. The approach does not attempt to model the full historical transmission dynamics since before MDA, which requires more complex models ^17^. We used a linear relationship between SCR and seroprevalence, *scr* = (π * 100)0.36, (see Methods). This empirical relationship was derived from a large number of surveys (n=48) ^8^, and should be robust for translation between seroprevalence and SCR for the same age range (1-5-years) and model (constant force of infection). Conversion of seroprevalence to SCR over different age ranges (e.g. 1-3-years or 1-9-years) or modelling assumptions (e.g. including seroreversion or immunity waning) would imply a different relationship that could be derived analytically or estimated empirically.

Our analyses could be extended to evaluate sampling strategies for integrated serology assessments, especially in conjunction with other neglected tropical diseases (NTDs). NTD programs are increasingly exploring integrated multi-pathogen serosurveillance as a cost-effective monitoring tool for endemicity mapping ^18–24^. Sampling efficiency could be optimized to embed a target sample size for trachoma in a larger multi-disease survey, while ensuring the sample meets the desired precision ^25^. Alternatively, multi-analyte serosurveillance for monitoring transmission of other diseases apart from NTDs and including vaccine-preventable diseases could be integrated into trachoma prevalence surveys. For trachoma, integrated serosurveillance designs might not always yield desired precision and power for SCR estimation, though in principle the designs evaluated here should translate in terms of clusters and number of children aged 1–5 years. Similarly, testing blood samples that arise through opportunistic research studies or passive surveillance among young children could also provide estimates of the SCR to assess the absence of trachoma, even if the estimates are not necessarily representative. Integrated serological surveys or less standardized serosurveillance could provide an initial signal to inform trachoma control programs however, consideration as to whether the resulting sample is sufficiently representative of the at-risk area is necessary before using the data for decision making ^20^. If SCR estimates from an integrated or opportunistic survey had insufficient precision with respect to the proposed thresholds, then programs or other disease control entities could consider a follow-up trachoma prevalence survey to measure other contextual factors or indicators (TF and *Ct* infection) for better decision-making.

There are other possible avenues to extend this work. Although our conclusions were largely invariant to the assumed cluster-level heterogeneity in seroprevalence (ICC = 0.002 versus 0.01), we could consider possible spatial heterogeneity of seroprevalence for improved surveillance, especially where the disease is highly focal and spatial clustering of high-infection communities has been observed ^26,27^. Geostatistical analyses including spatial or environmental covariates could improve the predictive power of the surveys where seroprevalence is spatially correlated ^28,29^, though there are several limitations to such analyses for trachoma decision-making and more work is needed before the routine use of geostatistical analyses is recommended for trachoma ^30^. Alternatively, geostatistical analyses might enable the same predictive power to be achieved from a smaller sample size ^31,32^. Costing and laboratory methods are the other important aspects to be considered for including serology in trachoma prevalence surveys. Adding serology measurements to trachoma prevalence surveys was found to be viable and cost-effective for supporting elimination efforts in the United Republic of Tanzania and Mozambique ^33^, and laboratory tests routinely used to measure antibodies to Pgp3 (multiplex bead assay (MBA), enzyme-linked immunosorbent assay (ELISA), and lateral flow assay (LFA)) have demonstrated high accuracy and precision across different settings ^34,35^.

In conclusion, we provide a range of feasible survey designs that a trachoma program could use to correctly classify an EU as being below or above specific SCR thresholds for programmatic actions. This new information should help trachoma programs incorporate serology measurements into the design of surveillance surveys. Inclusion of serology may be especially useful for assessing populations with low endemicity, including after validation of trachoma elimination as a public health problem. The results presented here will form the basis to further improve trachoma survey design, particularly the serology component.

## Data Availability

All data produced are available online through the Open Science Framework (https://osf.io/va8uc/ and https://osf.io/npsv9/)

## Global Trachoma Serosurveillance Study Group

Abdou Amza (Programme National de Lutte Contre la Cecité, Niamey, Niger), Abou-Bakr Sidik Domingo (Ministère de la Santé et de L’Hygiène Publique, Lomé, Togo), Adisu Abebe Dawed (Amhara Regional Health Bureau, Bahir Dar, Ethiopia), Ahmed M. Arzika (Centre de Recherche et d’interventions en Sante Publique, Niamey, Niger), Amir B. Kello (World Health Organization Regional Office for Africa, Brazzaville, Congo), Andres G. Lescano (School of Public Health and Administration, Universidad Peruana Cayetano Heredia, Lima, Peru), Ansumana Sillah (The National Eye Health Programme, Ministry of Health, Banjul, The Gambia,), Anthony W. Solomon (Global Neglected Tropical Diseases Programme, World Health Organization, Geneva, Switzerland), Ariktha Srivathsan (F.I. Proctor Foundation, University of California San Francisco, San Francisco, USA. Department of Epidemiology and Biostatistics, University of California San Francisco, San Francisco, USA), Beido Nassirou (Programme National de Lutte Contre la Cecité, Niamey, Niger), Belgesa E. Elshafie (Ministry of Health, Khartoum, Sudan), Benjamin F. Arnold (F.I. Proctor Foundation, University of California San Francisco, San Francisco, USA. Department of Ophthalmology, University of California San Francisco, San Francisco, USA. Institute for Global Health Sciences, University of California San Francisco, San Francisco, USA), Benjamin Marfo (Ghana Health Service, Accra, Ghana), Boubacar Kadri (Programme National de Lutte Contre la Cecité, Niamey, Niger), Catherine Oldenburg (F.I. Proctor Foundation, University of California San Francisco, San Francisco, USA. Department of Ophthalmology, University of California San Francisco, San Francisco, USA. Institute for Global Health Sciences, University of California San Francisco, San Francisco, USA.), Chris Drakeley (London School of Hygiene & Tropical Medicine, London, UK), Chrissy Roberts (London School of Hygiene & Tropical Medicine, London, UK), David Chaima (Kamuzu University of Health Sciences, Blantyre, Malawi), Diana L. Martin (Division of Parasitic Diseases and Malaria, Centers for Disease Control and Prevention, Atlanta, USA), Dionna M. Wittberg (F.I. Proctor Foundation, University of California San Francisco, San Francisco, USA), Dorothy Yeboah-Manu (Noguchi Memorial Institute for Medical Research, University of Ghana, Accra, Ghana), E. Brook Goodhew (Division of Parasitic Diseases and Malaria, Centers for Disease Control and Prevention, Atlanta, USA), E. Kelly Callahan (The Carter Center, Atlanta, USA), Emma M. Harding-Esch (London School of Hygiene & Tropical Medicine, London, UK), Emma Michelle Taylor (Sightsavers, Haywards Heath, UK), Eshetu Sata (The Carter Center, Addis Ababa, Ethiopia), Everlyn Kamau (F.I. Proctor Foundation, University of California San Francisco, San Francisco, USA), Fasihah Taleo (World Health Organization, Port Vila, Vanuatu), Fikre Seife (Federal Ministry of Health, Addis Ababa, Ethiopia), Harran Mkocha (Kongwa Trachoma Project, Kongwa, Tanzania), Jaouad Hammou (Direction de l’Epidemiologie et de Lutte contre les Maladies, Rabat, Morocco), Jeremy D. Keenan (F.I. Proctor Foundation, University of California San Francisco, San Francisco, USA. Department of Ophthalmology, University of California San Francisco, San Francisco, USA), John M. Nesemann (F.I. Proctor Foundation and Department of Ophthalmology, University of California San Francisco, San Francisco, USA), Karana Wickens (Division of Parasitic Diseases and Malaria, Centers for Disease Control and Prevention, Atlanta, USA), Katherine Gass (Neglected Tropical Diseases Support Center, Task Force for Global Health, Atlanta, USA), Khumbo Kalua (University of British Columbia, Vancouver, Canada. Blantyre Institute for Community Outreach, Blantyre, Malawi), Kimberly Fornace (Saw Swee Hock School of Public Health, National University of Singapore, Singapore, Singapore), Kristen Aiemjoy (Department of Public Health Sciences, School of Medicine, University of California Davis California, USA. Department of Microbiology and Immunology, Mahidol University Faculty of Tropical Medicine, Bangkok, Thailand), Kristen K. Renneker (International Trachoma Initiative, Atlanta, USA), Kwamy Togbey (Ministère de la Santé et de l’Hygiène Publique, Lomé, Togo), Laura Senyonjo (Sightsavers, Haywards Heath, UK), Mabula Kasubi (Muhimbili Medical Center, Dar es Salaam, Tanzania), Marcel S. Awoussi (Ministère de la Santé et de L’Hygiène Publique, Lome, Togo), Martha Idalí Saboyá Díaz (Department of Prevention, Control, and Elimination of Communicable Diseases, Pan American Health Organization, Washington DC, USA), Michael Peter Masika (National Eye Care Coordinator, Ministry of Health, Malawi), Mohammed Youbi (Epidemiology and Disease Control Directorate, Ministry of Health and Social Protection, Rabat, Morocco), Nishanth Parameswaran (Division of Parasitic Diseases and Malaria, Centers for Disease Control and Prevention, Atlanta, USA), Oliver Sokana (Solomon Islands Ministry of Health and Medical Services, Honiara, Solomon Islands), Patrick J. Lammie (Neglected Tropical Diseases Support Center, Task Force for Global Health, Atlanta, USA), Pearl Anne Ante-Testard (F.I. Proctor Foundation, University of California San Francisco, San Francisco, USA), PJ Hooper (International Trachoma Initiative, Atlanta, USA), Prudence Rymil (Ministry of Health, Port Vila, Vanuatu), Rabebe Tekeraoi (Ministry of Health and Medical Services, South Tarawa, Kiribati), Ramatou Maliki (The Carter Center, Niamey, Niger), Robert Butcher (London School of Hygiene & Tropical Medicine, London, UK), Robert Ko (University of Papua New Guinea, Port Moresby, Papua New Guinea), Robin L. Bailey (London School of Hygiene & Tropical Medicine, London, UK), Sarah Gwyn (Division of Parasitic Diseases and Malaria, Centers for Disease Control and Prevention, Atlanta, USA), Sarjo Kanyi (The National Eye Health Programme, Ministry of Health, Banjul, The Gambia), Scott D. Nash (The Carter Center, Atlanta, USA), Seth Blumberg (F.I. Proctor Foundation, University of California San Francisco, San Francisco, USA. Department of Ophthalmology, University of California San Francisco, San Francisco, USA. Institute for Global Health Sciences, University of California San Francisco, San Francisco, USA), Sheila K. West (Wilmer Eye Institute, Johns Hopkins University, Baltimore, USA), Solomon Aragie (F.I. Proctor Foundation, University of California San Francisco, San Francisco, USA. The Carter Center, Addis Ababa, Ethiopia), Stephanie J. Migchelsen (London School of Hygiene & Tropical Medicine, London, UK), Taye Zeru (Amhara Public Health Institute, Bahir Dar, Ethiopia), Thomas M. Lietman (F.I. Proctor Foundation, University of California San Francisco, San Francisco, USA. Department of Ophthalmology, University of California San Francisco, San Francisco, USA. Department of Epidemiology and Biostatistics, University of California San Francisco, San Francisco, USA. Institute for Global Health Sciences, University of California San Francisco, San Francisco, USA), Timothy William (Subang Jaya Medical Centre, Subang Jaya, Malaysia), Willie Pomat (PNG Institute of Medical Research, Port Moresby, Papua New Guinea), Zeinab Abdalla (The Carter Center, Khartoum, Sudan), Zerihun Tadesse (The Carter Center, Addis Ababa, Ethiopia)

## Data and materials availability

De-identified data and replication files required to conduct the analyses are available through the Open Science Framework (https://osf.io/va8uc/ and https://osf.io/npsv9/). Analyses used R statistical software (version 4.2.3).

## Author contributions

Following CRediT taxonomy: conceptualization (EK, BFA), data curation (all authors), formal analysis (EK, BFA), funding acquisition (AWS, SDN, BFA), methodology (EK, KG, EMH-E, MISD, PAA-T, KF, AWS, SDN,BFA), project administration (BFA), software (EK, BFA), supervision (BFA), validation (EK, BFA), visualization (EK, BFA), Writing – Original Draft Preparation (EK, BFA), Writing – Review & Editing (all authors).

## Ethics

The secondary analysis protocol was reviewed and approved by the Institutional Review Board at the University of California, San Francisco (Protocol #20-33198). All primary data that contributed to the analysis were collected after obtaining informed consent from all participants or their guardians under separate, local human subjects research protocols in accordance with the Declaration of Helsinki. Members from each contributing primary research study have participated as collaborators and co-authors on the present analyses from their initial stages, including the design, interpretation, and summary of results. Co-authors were nominated by each study’s principal investigator to represent the country and study teams that originally contributed the data. De-identified datasets made public through this analysis have been reviewed and approved by representatives from each study and conform with ethical guidelines set forth in the original protocols. Analyses were led by investigators at the University of California, San Francisco with guidance and input from all co-authors to incorporate local stakeholder perspectives.

